# Reward Responsiveness across Autism and Autistic Traits – Evidence from Neuronal, Autonomic, and Behavioural Levels

**DOI:** 10.1101/2022.02.11.22270801

**Authors:** Magdalena Matyjek, Mareike Bayer, Isabel Dziobek

## Abstract

Atypicalities in processing of social rewards have been suggested to lie at the root of social difficulties in autism spectrum conditions (ASC). While evidence for atypical reward function in ASC is mounting, it remains unclear whether it manifests specifically in hypo- or hyper-responsiveness, and whether it appears only in the social domain or more generally. Moreover, stimuli used as social rewards in studies often lack familiarity and relevance, which are known to enhance reward-related responses. In this study, we investigated behavioural (reaction times and ratings), neuronal (event-related potentials), and autonomic (pupil sizes) responses to three conditions – relevant social rewards, money, and neutral informative outcomes – in 26 ASC and 53 non-autistic subjects varying in levels of autistic traits, as measured with the Autism Spectrum Quotient (AQ). We used both a population-based approach (low AQ vs. high AQ) and a psychopathological approach (low AQ vs. ASC) to investigate the effects of both sub-clinical and clinical autistic traits on reward responsiveness. As hypothesised and preregistered, autism and autistic traits did not differently influence responses to social, monetary, and neutral outcomes on behavioural, neuronal or autonomic level. Although the ASC group rated the stimuli’s motivational and rewarding values lower than the other groups, the task performance was similar for all participants. Moreover, the ASC group in contrast to low AQ group showed enhanced brain responses (the CNV) in early anticipation and larger pupil constrictions in reward reception. Both effects were also predicted by autistic traits (AQ). Together, our results do not offer evidence for specifically social reward deficits in ASC. Instead, the data suggest enhanced neuronal and autonomic reward responsiveness linked to autism with simultaneously typical performance and reduced self-reported motivational and rewarding values of stimuli. Together, these results emphasise the need to investigate multiple processing levels for a broader picture of reward responsiveness in ASC.

## 1. Introduction

Rewards constitute a crucial factor in learning (Schultz, 2015) and thus are immensely important for life. The social motivation theory suggests that responsiveness to rewards in autism spectrum conditions (ASC) is atypical at least in the social domain (Chevallier et al., 2012). The consequence of this could be a cascade of neurodevelopmental difficulties including reduced pleasure from interacting with others, withdrawal from social situations, insufficient exposure to social stimuli, and finally social interaction difficulties. Thus, it was suggested that atypicalities in reward responsiveness may lie at the root of social difficulties in autism (Dawson et al., 2002; Dawson, Webb, Wijsman, et al., 2005).

Importantly, this account is based on observations that behavioural manifestations of altered social motivation (e.g., lower orienting towards social stimuli) are linked to reduced activation of the reward circuit, like the amygdala and the orbitofrontal cortex (Chevallier et al., 2012). Thus, hypo-responsiveness to rewards was speculated to be the underlying cause for the well-documented decreased behavioural motivation for social stimuli in autism (Chevallier et al., 2012). Such hypo-responsiveness has been previously quantified in multiple ways, e.g., hypoactivation of the reward circuit (Baumeister et al., 2020), diminished electrical brain activity (Kohls et al., 2011), decreased autonomic responses (Sepeta et al., 2012), or slower reactions (Demurie et al., 2011). The first formulations of the social motivation theory predicted that reward responsiveness is diminished in ASC especially in the social domain (Dawson et al., 2002; Dawson, Webb, & McPartland, 2005; Schultz, 2005) but more recent works suggest general atypicalities in this group, manifesting in both social and non-social domains (Bottini, 2018; Clements et al., 2018; Keifer et al., 2021; Kohls et al., 2012).

However, the empirical studies testing the predictions of the social motivation theory have yielded mixed results. While some published works report that ASC is related to hypo-responsiveness specifically to social (e.g., Scott-Van Zeeland et al., 2010; Sepeta et al., 2012; Stavropoulos & Carver, 2014b, 2014a) or both social and non-social rewards (e.g., Baumeister et al., 2020; Kohls et al., 2011, 2013; Kohls, Thönessen, et al., 2014; Richey et al., 2014), other studies found no differences between ASC and comparison groups (e.g., Demurie et al., 2016; Ewing et al., 2013; Gilbertson et al., 2017). Moreover, some studies reported *hyper*-responsiveness to rewards in ASC, especially to objects related to special interests (e.g., Cascio et al., 2014; Kohls et al., 2018; Watson et al., 2015), but also to other social and non-social rewards (e.g., Matyjek, Bayer, et al., 2020; Pankert et al., 2014; van Dongen et al., 2015). Similarly, a recent meta-analysis of neuroimaging studies found both hypo- and hyper-activations in the reward brain circuit in the ASC group in comparison to non-autistic individuals (Clements et al., 2018). However, while any difference between these groups may be considered atypical, both enhanced and attenuated reward-related responses are not sufficient or clear evidence to support the claims of the social motivation theory.

Although the social motivation theory has attracted considerable attention based on its attempt to explain autistic symptomatology, the heterogenous results have casted doubts on its validity. To interpret these mixed results, it is important to carefully consider the methodological aspects of experimental designs in the available studies (Bottini, 2018). For example, familiarity and social relevance of the stimuli (e.g., faces) used as social rewards might be important but rarely addressed aspects in processing of social rewards. Often, studies use a smiling face of an unknown person as a social reward stimulus; however, such an unknown face carries no relevance to the individual or in the current study situation. In contrast to unknown faces, familiar and relevant faces are rated as more rewarding and elicit higher activation in reward-related brain structures (Acevedo et al., 2012; Bayer et al., 2021; Matyjek et al., 2021; Sugiura, 2014). Moreover, familiarity of faces has the potential to improve otherwise atypical face processing in ASC (Pierce et al., 2004; Pierce & Redcay, 2008). For these reasons, as social rewards in this study, we used pictures of the smiling face of the main experimenter, who is a familiar and relevant person in the study context (also see Hayward et al., 2018; Matyjek, Bayer, et al., 2020). Additionally, when feedback in an experiment is delivered with a picture of a face, it is important to address social anxiety traits in the participants, as anxious individuals are especially sensitive to social evaluation (Spain et al., 2018). Moreover, social anxiety often co-occurs with autism (Bejerot et al., 2014; Bellini, 2006) and has been previously linked to atypical reward processing (Cremers et al., 2015; Richey et al., 2014). Therefore, we planned to control for the modulatory effects of social anxiety traits on reward responsiveness.

Further, even though autistic traits in the general population have been repeatedly related to atypicalities in reward processing (Carter Leno et al., 2016; Cox et al., 2015; Dubey et al., 2015; Matyjek et al., 2020; Rolison et al., 2017; Sims et al., 2013), studies contrasting ASC and non-autistic individuals rarely control for autistic traits in the latter. Importantly, autistic traits are distributed normally across the general population (Baron-Cohen et al., 2001; Hoekstra et al., 2007; Ruzich et al., 2015), they are aetiologically linked to autistic traits in ASC, and they seem to assess the same latent constructs in ASC and non-clinical sample. Thus, studying effects of autistic traits on reward responsiveness in subclinical populations could assist in identifying relevant phenomena for ASC. Moreover, at least some of the inconsistencies in the literature investigating reward processing in autism may be due to the level of autistic traits in the comparison groups, as neglecting them may render it difficult to compare group effects between studies. Therefore, to provide a broader picture of reward processing in the autism spectrum, in the current study we investigated it using both *population-based* and *psychopathological* approaches, i.e., we compared individuals with low levels of autistic traits (and no autism diagnosis) to those with high levels of the traits (and no diagnosis), and to individuals diagnosed with autism.

Finally, to interpret the mixed results in the literature, neuronal atypicalities should be linked to behavioural manifestations of social difficulties in ASC (yet, evidence for this link is often lacking; Baumeister et al., 2020; Kohls et al., 2013; Scott-Van Zeeland et al., 2010). Capturing indexes of reward responsiveness on multiple levels has the potential to inform the interpretation of conflicting results in the literature and provides a more complete picture of the process. For example, behavioural indexes of an investigated process might aid the interpretation of co-recorded neuronal activity: Studies investigating fusiform activation in response to observing faces in ASC have yielded mixed results, but these inconsistencies may be explained by task demands, as shown by a negative correlation between the activation in this area and behavioural performance (Shafritz et al., 2015). Similarly, neuronal and autonomic measures have been shown to contribute differently to decision making, perceptual discrimination, and interoceptive attention (Al et al., 2020; de Gee et al., 2017; Waschke et al., 2019). Although several indexes of reward responsiveness have been investigated in the past research in the context of autism, including behavioural (e.g., reaction times, effort, accuracy; Demurie et al., 2011; Ewing et al., 2013; Geurts et al., 2008), neuronal (neuroimaging and electroencephalography (EEG); e.g., Kohls et al., 2011; Scott-Van Zeeland et al., 2010), and autonomic (e.g., electrodermal activity, pupil sizes; Neuhaus et al., 2015; Sepeta et al., 2012) levels, to our knowledge, no study to date has collected responses from all three levels from the same sample in a reward-related paradigm. In the current study, we fill this gap by reporting behavioural indexes of reward responsiveness (ratings of motivational and rewarding values of stimuli as well as combined measures of reaction times and accuracy), event-related potentials (ERPs), which offer excellent temporal resolution allowing for separate estimations of reward processing phases: anticipation and reception (Berridge, 1996, 2009; Berridge et al., 2009), and pupillary responses, which reflect the neuronal activation in the locus coeruleus (LC), a structure vastly involved in reward processing and motivation (Aston-Jones et al., 1999; Bast et al., 2018; Bouret & Richmond, 2015).

This study set out to investigate behavioural, neuronal, and autonomic responses in anticipation and reception of monetary and relevant social rewards as well as neutral outcomes across individuals with different levels of autistic traits and with autism. In our modified design with a socially relevant context, we expected that the familiar social stimuli would enhance reward-related responses in individuals with high levels of autistic traits and autism. Thus, on all processing levels (neuronal, autonomic, and behavioural), we expected to observe similar effects during anticipation and reception of social, monetary, and neutral incentives across groups and autistic traits (Barman et al., 2015; Demurie et al., 2013, 2016; Ewing et al., 2013; Gilbertson et al., 2017; Neuhaus et al., 2015). Further, conforming to the results from a similar study design of our group (Matyjek et al., 2020) and in line with work of other groups (Pankert et al., 2014; van Dongen et al., 2015), we expected to observe enhanced neuronal and autonomic responses in individuals with ASC and high levels of autistic traits in contrast to those with low trait levels. Because the available research suggests that ASC-specific atypicalities in reward responsiveness are more pronounced in the anticipation than reception phase (Keifer et al., 2021; Kohls et al., 2012), and our previous results identified differences between early and late phases of reward anticipation, we further expected that these group differences would be stronger in early than in late anticipation (Matyjek et al., 2020), but would not be observed in reception (Bottini, 2018). Finally, in order to confirm that the targeted responses are reward-related, we predicted to see larger responses to rewarded conditions (social and monetary) in all measures, as compared to the neutral outcomes (Cox et al., 2015; Kohls et al., 2011).

In addition to testing these primary hypotheses, we aimed to explore several secondary analyses. First, to further quantify behavioural indexes of reward responsiveness, we collected scores estimating inhibition and approach tendencies from the participants and aimed to relate them to the neuronal and autonomic measures as well as autistic traits. Second, although we were primarily interested in *reward* responsiveness and for that the primary analyses were conducted on the data from successful trials (where reward could be obtained), we also explored the neuronal and autonomic responses in the reception of unsuccessful (non-rewarded) trials. Finally, for a dimensional analysis of autistic traits (instead of group-based), we explored whether the trait levels across all participants predict the reward-related reaction times, ERPs, and pupillary responses in linear and non-linear models.

## 2. Methods

The methods, hypotheses, and analyses were preregistered at https://osf.io/3re72. Data, analysis code in R, and an html file including all analyses steps and results can be found at https://osf.io/vse38/.

### 2.1 Sample size determination

To estimate the sample size, we performed a power analysis with the g*power software (Faul et al., 2009), with power set to 0.8 and with an intermediate effect size f = 0.302. The effect size was calculated from the between-subject factor of group (high vs. low autistic traits) in response to reward cues in our previous experiment (Matyjek et al., 2020). This analysis yielded a total sample size of 52 with 26 data sets in each of two groups planned for comparisons (ASC vs. low autistic traits, and high vs. low autistic traits). Based on this, we planned to recruit 26 participants per group, summing up to the total of 78 participants.

### 2.2 Participants

A total of 82 volunteers across three groups (ASC, low- and high autistic traits) participated in the study. The data sets of 3 participants (two from the ASC group) were excluded due to poor EEG signal quality (1), refusal to perform a task involving money (1), and a technical issue with EEG recording (1). Demographic information for all groups with group comparisons are summarised in Table 1. All participants provided written informed consent; the study was approved by the ethics committee of the Faculty of Psychology of the Humboldt-Universität zu Berlin and was conducted in accordance with the Declaration of Helsinki. After the experiment, the aims of the study were revealed to the participants in a debriefing conversation. Participants were compensated 8 Euro per hour plus additional 4 Euro as a monetary reward earned during the task (for details, see section 2.3), which resulted in a total of 30-40 Euro.

**Table 1.** Demographic and trait characteristics of subject samples in all groups. Count is provided for gender and means (with standard deviations) for all other items. HAQ = high autistic traits, LAQ = low autistic traits, ASC = Autism Spectrum Disorder, AQ = Autism Spectrum Quotient score, BIS/BAS = Behavioural Inhibition/Approach System, LSAS-SR = Liebowitz Social Anxiety Scale, R:N/L = Right, Neutral/Left (handedness), OR = odds ratio in Fisher’s Exact Test. Statistically significant tests were marked with ** for p < .01 and *** for p < .001.

| Group description |  |  |  |
| --- | --- | --- | --- |
|  | LAQ<br>N = 26 | HAQ<br>N = 27 | ASC<br>N = 26 |
| Gender – female:male | 18:8 | 12:15 | 13:13 |
| Age (years) | 30.6 (7.9) | 31.9 (10.6) | 32.7 (10.7) |
| AQ (total) | 10.9 (2.3) | 25.9 (5.5) | 39.3 (5) |
| BAS drive | 12.5 (2.1) | 11.6 (1.9) | 11.2 (2) |
| BAS reward responsiv. | 16.8 (1.7) | 15.7 (2.3) | 14.9 (2.3) |
| BAS fun seeking | 12.5 (1.8) | 11.2 (1.9) | 9.7 (2.7) |
| BIS | 18.8 (3.2) | 20.9 (4.7) | 23.5 (3.8) |
| LSAS-SR | 31.1 (15.9) | 58.6 (27.2) | 83.5 (32.2) |
| Handedness – R:N/L | 25:1 | 25:2 | 20:6 |
| Between-group differences |  |  |  |
|  | LAQ vs. HAQ | LAQ vs. ASC |  |
| Gender | OR = 2.76 | OR = 2.21 |  |
| Age (years) | $t = -0.51$ | $t = -0.79$ | |
| AQ (total) | $t = -13.02^{***}$ | $t = -26.25^{***}$ | |
| BAS drive | $t = 1.71$ | $t = 2.35^*$ | |
| BAS reward respons. | $t = 1.87$ | $t = 3.42^{***}$ | |
| BAS fun seeking | $t = 2.68^*$ | $t = 4.43^{***}$ | |
| BIS | $t = -1.93$ | $t = -4.93^{***}$ | |
| LSAS-SR | $t = -4.51^{***}$ | $t = -7.42^{***}$ | |
| Handedness | OR = 0.51 | OR = 6.25 |  |

#### 2.2.1 Non-autistic participants

Non-autistic participants were recruited via internet advertising platforms and flyers distributed at Berlin’s university campuses. Inclusion criteria were age (18-50), proficiency in German, no history of psychological, neurological, or psychiatric disorders in the last 6 months (including medication), and no past diagnosis of such. Interested volunteers were asked to complete the Autism Spectrum Questionnaire (AQ; Baron-Cohen, Wheelwright, Skinner, et al., 2001) and were invited to participate based on the score (we aimed to increase the spread of the scores and to balance the size of low and high scoring groups). The mean AQ score in the non-autistic group (*N* = 53) was 18.6 (*SD* = 8.7); groups with high (HAQ) and low (LAQ) autistic traits (*N* = 25 and 26, respectively) were created based on a median split (*Mdn* = 17). This sample (30 females and 23 males) had a mean age of 31.3 (*SD* = 9.3). All participants had normal or corrected-to-normal vision and 50 were right-handed (based on the Edinburgh Inventory, Oldfield, 1971). One participant reported attending a psychotherapy in the last six months, and two earlier than that. No participants in this group had been medicated with psychopharmaceuticals.

#### 2.2.2 ASC group

Participants with ASC were recruited via a specialised autism outpatient clinic at Charité— Universitätsmedizin Berlin, and the Outpatient Clinic for Social Interaction, University Outpatient Clinic for Psychotherapy and Psychodiagnostics of Humboldt-Universität zu Berlin, an online forum for the autism community (www.aspies.de), and internet advertising platforms. All participants were confirmed to have a prior diagnosis matching the DSM-5 criteria for autism spectrum disorder (American Psychiatric Association, 2013) (22 participants were diagnosed with Asperger’s Disorder, 1 participant was diagnosed with Atypical Autism, and for three subjects only the information was available that they were diagnosed with an autism spectrum disorder) made by professionals in specialised autism-diagnosis centres (the diagnosis was confirmed directly by the centres and/or by a written diagnosis provided by the participants). In 20 cases, data were available for the Autism Diagnostic Observation Schedule (ADOS; Lord et al., 2000), and in 17 also the Autism Diagnostic Interview-Revised (ADI-R; Bölte & Poustka, 2001; Lord et al., 1994), a semi-structured interview administered to the caretakers. Additionally, inclusion criteria were age (18-50) and proficiency in German. All participants had normal or corrected-to-normal vision and 20 participants were right-handed. Several participants in the ASC group reported comorbid psychopathology and/or receiving psychotherapy in the last six months (N=6), and earlier (N=5) (all for depression and/or anxiety). Four participants were medicated at the time of the study or in the last six months and two more earlier than that (all with selective serotonin or serotonin-norepinephrine reuptake inhibitors).

### 2.3 Stimuli and task

We adapted a cued incentive delay task (Knutson et al., 2000) to include both social and non-social rewards. Participants were shown a cue indicating a possible reward in a given trial and asked to speedily respond to a following target. For correct responses they received either a reward – a picture of the experimenter’s smiling face (social reward) or money (5 cents, non-social reward), or a reward-neutral outcome (an informative letter). The instructions were delivered both in writing and verbally.

Each trial started with a cue presented at the centre of the screen for 1500 ms, which indicated the condition. Three conditions were introduced: social (S), monetary (M), and neutral (N). The cues consisted of white letters on black squared background (4 × 4° of visual angle). For clarity, the letters were linked to the names of potential outcomes in German: “L” for *Lächeln* (smile), “M” for *Münze* (coin), and “N” for *Neutral*. The cue was followed by a small white fixation cross (30 × 30 pixels) displayed centrally for 500 ms. Then, a blue or a purple target (a circle, 1 × 1° of visual angle) was displayed in the centre. The display time of the targets was adapting to each participant’s performance: Every four trials the display time was increased by 20% if in the last four trials no more than 1 response was correct, kept the same if 2 responses were correct, or decreased by 20% when 3 or 4 responses were correct. This procedure ensured approx. 60% accuracy on average across all participants. Upon detection of the target’s colour, participants were required to press one of two response buttons as fast as possible. The colour of the target in each trial was random and the response keys corresponding to the colours were counterbalanced across participants. A trial was successful when participants pressed the correct button during the display of the target. After the button press (or the end of the display time, in case of missing responses), the pre-feedback waiting period with a fixation cross was presented for 1500 to 2000 ms (jittered across trials). Then, a feedback stimulus (matching the incentive type indicated by the cue) was presented in the centre of the screen for 2000 ms.

Correct responses in the S condition were rewarded with a picture of a happy/approving face of the main experimenter. In the M condition, a picture of a “5” coin was presented, and in the N condition, letter “R” as an indicator of successful response (for German *richtig* meaning *correct*). Incorrect responses in S, M, and N conditions were followed by a face with a neutral expression, a “0” coin, and letter “F” (for *falsch*, or *wrong*). Letters in N and coins in M were displayed on a background made of scrambled pixels of the S rewarding feedback picture (the happy/approving face). All feedback stimuli were equal in size (4 × 4° of visual angle) and luminance (ensured with the mean value of luminance in perceptual space in GIMP 2.0, which was additionally confirmed with a photometer). Participants were instructed that a “5” coin meant they were receiving additional 5 cents. The current balance was displayed after each block.

Each of the six blocks consisted of one condition. The blocks were presented pseudo-randomly: The first three blocks were presented in random order and the last three blocks repeated that order. Each block consisted of 50 trials, resulting in a total of 300 trials. Before the start of the actual experiment, three blocks (one for each condition) of 10 trials were presented as training.

### 2.4 Procedure and socialising

After signing the consent form, the participants were prepared for the EEG recording, which took ca. 20 min. During this time the experimenter had a light social conversation following a semi-scripted interaction. The aim was to provide a natural acquaintance with the experimenter, with whom all participants spent the same amount of time. Moreover, this allowed the participants to familiarise with the experimenter’s face in a natural fashion: from various angles and with various facial expressions. To emphasize the shared social context, the experimenter also indicated that this research was her project, and that she appreciated the subjects’ participation in the study. Then, participants were seated in an electrically shielded room at a distance of 70 cm from a 19-inch computer screen and 85 Hz refresh rate. To keep the light conditions constant, the room was artificially lit. Participants were asked to place their chin and forehead on a headrest in order to restrain movements. The experiment was programmed and executed in MATLAB. Task instructions were displayed on the screen and additionally repeated verbally by the experimenter. Participants were asked to identify the person on the pictures used in the S condition prior to the training and all correctly recognised the experimenter. After the recording, participants answered a number of debriefing questions on a computer screen. In the end they were debriefed and informed about in details about the purpose of the study.

### 2.5 Behavioural measurements

#### 2.5.1 Reaction times

As a measure of performance, we collected participants’ reaction times in successful trials (those which ended in positive feedback, i.e., the correct button was pressed during the display time of the target) in the task. Both shorter reaction times and higher accuracy have been previously interpreted as indexes of increased motivational and reinforcing values of the related rewards (e.g., Neuhaus et al., 2015). However, since faster responses may lead to lower accuracy, increasing response time to ensure more successful trials (and thus rewards) may be used as a strategy. Therefore, as a behavioural index of reward processing, we targeted reaction times (excluding those faster or slower than two standard deviations from a subject’s mean) corrected for accuracy, i.e., the linear integrated speed-accuracy score (LISAS; Vandierendonck, 2018).

#### 2.5.2 Debriefing questions

Further, we collected self-reported measures of reward responsiveness. Participants answered the following debriefing questions: *How motivated were you in the experiment?* (general motivation); *How important was the reward type to you?* (importance of condition); *How often, right after giving the response, did you feel you knew whether you were successful?* (sense of agency), *How motivating did you find the cues?* (motivational value of cues); *How rewarding did you find the feedback pictures?* (rewarding value of feedback).

#### 2.5.3 Questionnaires

Finally, we administered several questionnaires. To quantify autistic traits, we used the Autism Spectrum Quotient (AQ; Baron-Cohen, Wheelwright, Skinner, et al., 2001) and the Social Responsiveness Scale (SRS; Constantino & Gruber, 2005), which were significantly correlated in our sample, *r*(65) = .66, *p* < .001. While AQ was our primary measurement of autistic traits, we also planned to explore how the SRS relates to other measures of reward responsiveness. However, fourteen participants did not feel comfortable asking a close person to fill the SRS questionnaire for them (in contrast to the AQ which is a self-administered tool, the SRS is completed by another person). Given that the SRS scores were only available for 65 participants (7 missing in ASC, 4 in HAQ, and 3 in LAQ), we did not focus on these data any further.

To quantify further reward-related behaviour, participants were asked to fill the Behavioural Inhibition and Approach Systems Scale (BIS/BAS; Carver & White, 1994). This questionnaire assesses the behavioural activation system (BAS) responsible for increased motivation and positive affect in response to incentives, and the behavioural inhibition system (BIS), linked to experiences of anxiety, fear, and negative affect in response to threatening stimuli. BAS is further divided into three subscales: drive (inclination to pursue desired goals); fun seeking (desire for new rewards); and reward responsiveness.

Finally, to control for social anxiety traits in all statistical models, we used the Liebowitz Social Anxiety Scale self-reported (LSAS-SR; Liebowitz, 1987), which assesses anxiety related to experiencing everyday social situations.

For each questionnaire, higher scores are interpreted as higher expressions of the targeted behaviour or trait. The mean scores for each group are presented in Table 1.

### 2.6 EEG data acquisition and pre-processing

For reward reception, we quantified the P3, which is a positive potential peaking around 300 ms after stimulus onset and reflecting an elaborated cognitive and affective function linked to reward (Wu & Zhou, 2009). Based on our previous research, we divided reward anticipation into early and late phases (Matyjek et al., 2020). For those, respectively, we targeted the Contingent Negativity Variation (CNV) and the Stimulus Preceding Negativity (SPN). These ERPs are slow negative waves peaking before a signal stimulus triggering a prompt action (CNV) or before receiving stimuli carrying important information, like feedback (SPN). Both CNV and SPN are linked to motivation and effort, and they reach higher amplitudes for anticipated affective or emotionally salient stimuli (Broyd et al., 2012; Brunia et al., 2012). As such, they have previously been used as indexes of reward anticipation (Matyjek et al., 2020; Stavropoulos & Carver, 2014b, 2014a).

The continuous EEG signal was recorded from 64 silver/silver-chloride active scalp electrodes (Biosemi Active Two) at a sampling rate of 512 Hz. An elastic cap with the extended 10-20 international electrode placement system was used. The collected signals were referenced online to the CMS-DRL ground loop, which drives the average potential as close as possible to the amplifier zero. The electrode offsets were kept within the range of ± 20 µV. Six external electrodes were used: four electrodes were placed at the outer canthi and below the eyes (to collect the horizontal and vertical electro-oculograms) and two were placed on the mastoids. Two online filters were applied: a 100 Hz low-pass and 0.01 Hz high-pass.

The offline pre-processing steps were performed using BrainVision Analyzer (Brain Products GmbH, Munich, Germany), in which all signals were re-referenced to average reference and filtered with a low-pass filter of 40 Hz (slope 8 dB/oct). The continuous data were segmented into segments ranging from -100 ms before to 7500 ms after the cue onset. A pre-cue baseline of 100 ms was applied. To identify and remove blinks and eye movements, an independent component analysis algorithm (restricted fast ICA) was used. Channels with low quality and noisy signals were interpolated using spherical splines of order 4 (2.7 % of all channels). Further, to exclude artifacts, a semi-automatic procedure was applied targeting signals exceeding ± 100 µV or voltage steps larger than 100 µV. This led to rejection of 5.39% of trials for all cue signals, 5.9% of trials for all pre-feedback, and 5.98% for all feedback signals. The data were divided into three sub-segments representing the phases: the incentive cue (early anticipation), the pre-feedback (late anticipation), and the feedback (reception). Those were ranging, respectively: from -100 ms before to 1500 ms after the cue onset; from -600 ms before feedback onset to feedback onset; and from -100 ms to 2000 ms after the feedback stimuli onset.

Across participants an average of 95 artifact-free successful trials was obtained in conditions M and S and 94 in N for the cue responses (*SD*_*M*_ = 5.83, *SD*_*S*_ = 6.08, *SD*_*N*_ = 8.11). In pre-feedback and feedback, the average number of successful trials was 57 (*SDpre-feedback* = 5.67, *SDfeedback* = 5.88). Average number of segments respectively for N, M, and S were in pre-feedback: 55.66 (6.37), 58.66 (4.86), and 57.19 (5.34), and in feedback: 55.46 (6.89), 58.61 (4.87), and 57.18 (5.34). The number of artifact-free segments did not vary significantly between conditions (N, M, S) in either of the phases (all *F* < 1.17, *p* > .31).

The temporal windows and regions of interest for the brain responses were chosen based on prior research and visual inspection of grand averages. For the early and late anticipation phases the time windows for the anticipatory brain responses were respectively the CNV and the SPN defined as the last 500 ms of each phase: in early anticipation this was 1000 – 1500 ms after cue onset (with -100 – 0 ms baseline), and in the jittered late anticipation phase this was -500 – 0 ms time-locked to feedback onset (with -100 – 0 ms baseline locked to the onset of the pre-feedback fixation cross). In the reception, the P3 was identified from 230 to 500 ms after feedback onset. Mean amplitudes for these time windows were calculated from electrodes Pz, P1, P2, POz, PO3, PO4, Oz, O1, O2. Finally, the mean amplitudes of the CNV, SPN, and P3 were aggregated per participant and condition and used in the statistical analyses.

### 2.7 Pupillary data acquisition and pre-processing

We recorded the pupil sizes in both anticipation and reception phases of reward processing. The pupil has been observed to increase in size while anticipating rewards (Cash-Padgett et al., 2018; Koelewijn et al., 2018; Schneider et al., 2018; Takarada & Nozaki, 2017). In contrast, when receiving and evaluating outcomes, the pupil is negatively correlated with their reward values (Cash-Padgett et al., 2018; Matyjek, Bayer, et al., 2021; our pre-registered expectations of dilations in this phase were yet uninformed by this recent research). The contrasting pupillary responses in reward anticipation (stronger dilation for larger rewards) and reception (stronger constriction for larger rewards) emphasise that these phases are not a unitary construct and have qualitatively different elements (Cash-Padgett et al., 2018).s

Pupillary responses were recorded binocularly with a desktop-mounted eye tracker (Eye Tribe, TheEyeTribe) with a 60 Hz sampling rate. The EyeTribe Toolbox for Matlab (https://github.com/esdalmaijer/EyeTribe-Toolbox-for-Matlab) was used to send event triggers. The calibration was conducted with a nine-point grid and accepted when accuracy of < 0.7 degree was achieved. Data sets with poor data quality (more than 50% missing trials, with a trial removed when missing over 50% samples) were excluded from further processing and analysis (13). Offline preprocessing was performed with Matlab code published by Kret & Sjak-Shie (2018) with their default settings (upsampling was reduced from 1000 to 100 Hz). This includes blink and missing data interpolation, filtering and smoothing. Then, using a custom code in R ver. 4.0.2 (R Core Team, 2020), segmentation of the signal into phases with a subtractive baseline correction was performed: -200 to 0 ms before the cue onset for early anticipation, and -200 to 0 ms before the pre-feedback for late anticipation and for reception. Finally, the mean pupil size was calculated for each segment: 0 to 1500 ms after cue onset (early anticipation), -1500 to 0 ms before feedback onset (late anticipation), and 0 to 2000 ms after feedback onset (reception) and aggregated for participants and conditions.

In the remaining 66 datasets (N in LAQ, HAQ, and ASC was 23, 24, and 19, respectively), on average 87 trials per condition in the cue signals entered analyses (*SD*_*M*_ = 13.13, *SD*_*N*_ = 15.45, *SD*_*S*_ = 13.19). In pre-feedback and feedback, average number of only successful trials for M, N, and S conditions were, respectively, 55, 53, and 53 (pre-feedback: *SD*_*M*_ = 8.27, *SD*_*N*_ = 9.77, *SD*_*S*_ = 7.96, feedback: *SD*_*M*_ = 8.27, *SD*_*N*_ = 9.76, *SD*_*S*_ = 7.96). The number of trials did not differ between conditions in the cue signals, *F*(2,130) = 0.07, *p* = .936. It differed in pre-feedback, *F*(2,130) = 3.13, *p* = .047, and feedback, *F*(2,130) = 3.11, *p* = .048, but no contrasts survived corrections for multiple comparisons (all *p*_*corr*_ >= .075).

### 2.8 Data analysis

All data analyses were performed using R ver. 3.4.3 (R Core Team, 2020). The significance level for all the tests was set to .05. The data and analysis code (as well as an html file presenting all the analyses (primary and secondary) in an accessible way without the need to run the code) are available in the OSF repository: https://osf.io/vse38/.

#### 2.8.1 Primary analyses

As registered, we analysed reward-related ERPs, pupil sizes, and reaction times corrected for accuracy in two approaches: 1) *population-based approach*, which includes individuals with high levels of autistic traits as compared to individuals with low levels of autistic traits (groups created based on AQ score median split); and 2) *psychopathological approach*, which includes individuals diagnosed with autism as compared to individuals with low trait levels. Participants’ responses to the debriefing questions were analysed across the three groups, with Pearson’s correlation or linear models.

For reaction times, brain, and pupillary responses, we built multiple regression models with mixed effects (random intercepts for subjects) with the lmerTest package ver. 3.1-2 (Kuznetsova et al., 2017). Regression assumptions were checked and met for all models (normality, linearity, multicollinearity, homoscedasticity). Models, which violated these assumptions, were considered for outliers (based on influence and deletion diagnostics). Models with outliers were re-fitted after either overwriting a data point with the group’s mean in the given condition, or exclusion of a subject’s data set. Since condition is a multilevel categorical predictor (N, S, M), for the estimation of its main effect an analysis of variance (ANOVA) with Satterthwaite approximation for degrees of freedom was calculated on the models. For all reported post-hoc tests, a Holm correction was applied. Because there is no established way of calculating standardised effect sizes for mixed models’ terms (Rights & Sterba, 2018), the unstandardised slope estimates can be treated as effect sizes (Pek & Flora, 2018). However, to comply to the convention, as an approximation we also calculated partial Cohen’s *f* (*f*_*p*_) from ANOVAs performed on the models with the effectsize package ver. 0.3.2 (Ben-Shachar et al., 2020). As registered, we controlled for social anxiety traits by including the (centred) LSAS-SR score in all the models. Overall, all the models were built in the following form:

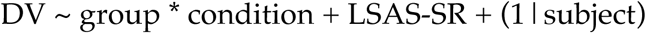

where DV is the dependent variable (LISAS, ERP (CNV, SPN, P3), or pupil size), and the term (1|subject) adds a random intercept for each subject. Because our hypothesis was that groups would respond similarly to different conditions (i.e., no interaction of group and condition), we first checked whether the interaction term was statistically significant. Since an insignificant effect does not mean a true negative effect, we used Bayes factors (BF) to provide an explicit quantification of evidence in favour of a model without the interaction vis-à-vis a model with the interaction (van Ravenzwaaij et al., 2019). In case of strong evidence in favour of a model without the interaction term, we continued the analysis with the model including only main effects.

#### 2.8.2 Secondary analyses

Methods and results for all secondary analyses are presented in the supplementary material. As registered, we explored (1) correlations between questionnaires (AQ, BIS/BAS, and LSAS-SR), ERPs, and pupil sizes, and (2) reaction times, ERPs, and pupillary responses also in unsuccessful (not rewarded) trials. Additionally to the registered analyses, we performed (3) dimensional analyses of AQ as a predictor of reward-related responses across all participants, and (4) exploratory analyses of the effects of age and gender in all the primary models.

## 3. Results

### 3.1 Primary analyses

The grand averages of the brain and pupil responses for all groups are shown in Figure 1 and Figure 2. Mean responses in both measures across conditions, groups, and phases are shown in Table 1 in the supplementary material. All analysis steps are shown in the html file (https://osf.io/vse38/) in sections 5.1.6., 5.2., and 5.3. for the population-based approach, and sections 6.1.6., 6.2., and 6.3. for the psychopathological approach.

**Figure 1.**
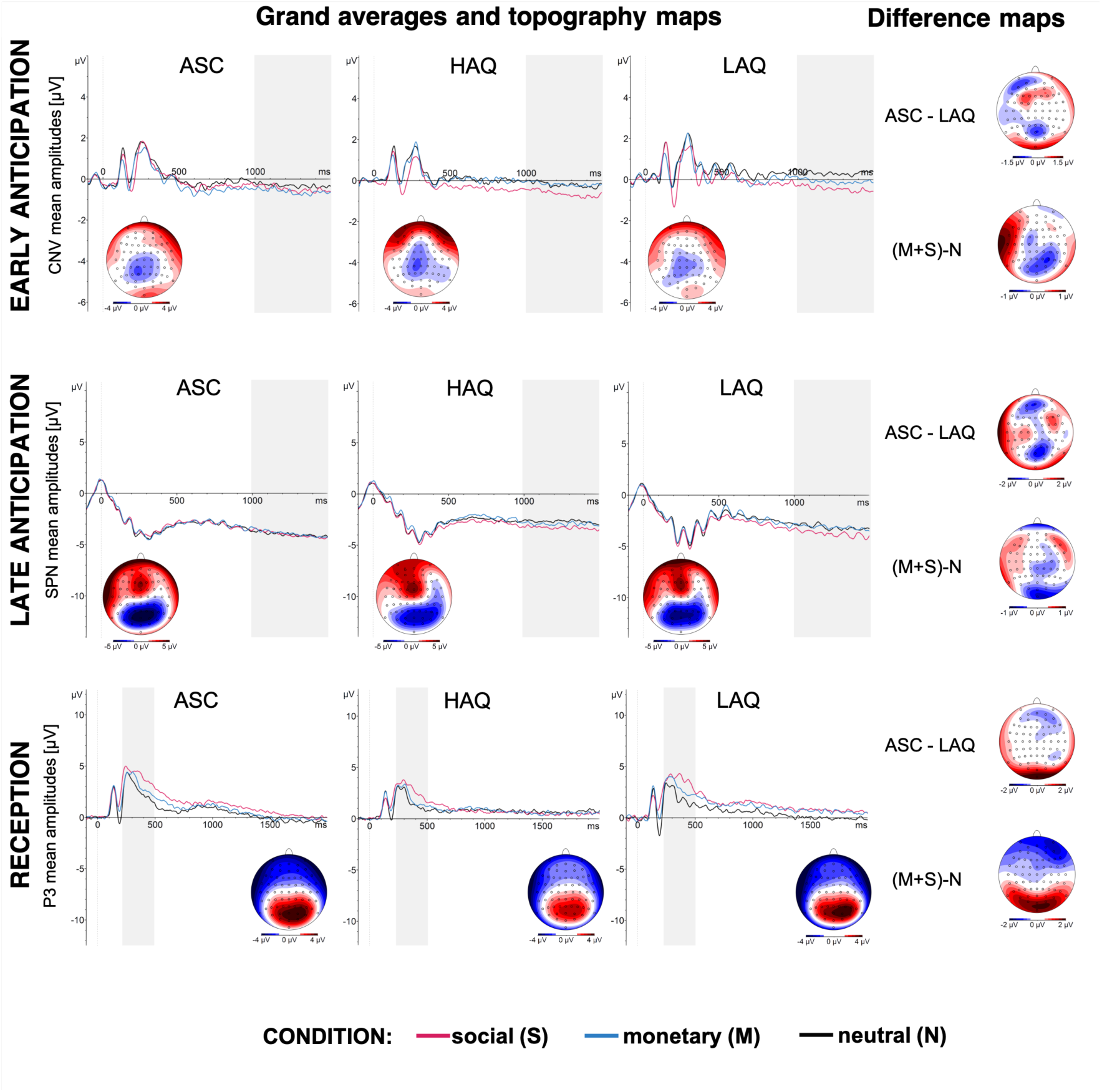
Grand averages of ERPs. The panels show the grand averages for groups (low autistic traits, LAQ; high autistic traits, HAQ; and autism, ASC) and conditions (social, S; monetary, M; and neutral, N) for each reward processing phase: early anticipation (top panel), late anticipation (middle panel), and reception (bottom panel). The dotted vertical lines mark the onset of each phase: cue presentation in early anticipation, fixation cross in late anticipation, and feedback in reception. The grey rectangles mark the time window for analyses. Note that for the purpose of visualisation, the SPN in the late reception is plotted as aligned to the onset of the fixation cross until 1500 ms, even though the display time was jittered (1500 – 2000 ms). The analysis included the last 500 ms before the feedback in each trial.

**Figure 2.**
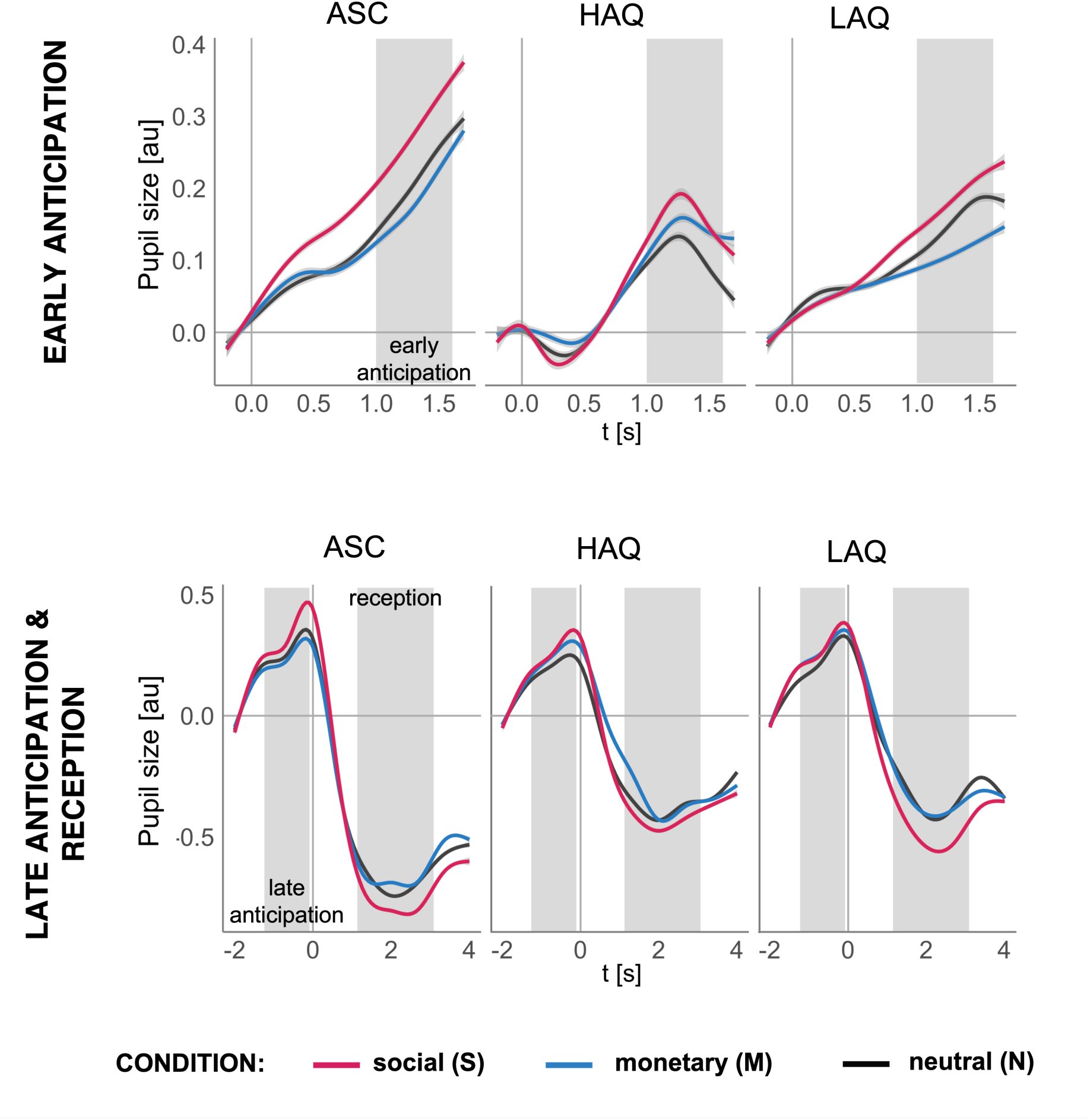
Grand averages of pupillary responses. The panels show the grand averages for groups (low autistic traits, LAQ; high autistic traits, HAQ; and autism, ASC) and conditions (social, monetary, and neutral) for each reward processing phase: early anticipation (top panel), late anticipation (middle panel), and reception (bottom panel). Stimuli shown in each phase and condition are displayed on the right side. The plots were created with generalized additive model smoothing and the grey shades show 95% confidence interval of this fit. The grey shaded areas mark time windows used for analyses. For visualisation purposes in the early anticipation and in the reception, respectively, the following 500 ms of fixation cross and 1000 ms of the intertrial interval were plotted. In the joint plot for late anticipation and reception 0 marks the onset of the feedback.

As predicted, in none of the models (across all measures and approaches) we observed an interaction effect of group (LAQ, HAQ, ASC) and condition (S, M, N), all *F* <= 2.09, all *p* >= .13. Moreover, in all analysis we found strong evidence in favour of models without the interaction term (all *BF* >= 20; Kass & Raftery, 1995) and those models showed a better fit (based on BIC) than models including this term. Hence, in the following we report only results of models re-fitted without the interaction term. Nevertheless, all analyses steps can be found in the code and the html file.

#### 3.1.1 Population approach (low AQ vs. high AQ)

##### 3.1.1.1 Reaction times

The factor condition significantly predicted reaction times corrected for accuracy, i.e., LISAS scores, *F*(2,100) = 11.7, *p* < .001, *f*_*p*_ = 0.48, with fasted responses in M than N (*p*_*corr*_ < .001, *est* = 9.99) and M than S (*p*_*corr*_ = .001, *est* = 7.37). Group was not a significant predictor (*f*_*p*_ = 0.13). For details and additional analyses of uncorrected reaction times and accuracy, see the html file.

##### 3.1.1.2 ERPs

Analyses of the CNV in the early anticipation, the SPN in the late anticipation, and the P3 in the reception all yielded a main effects of condition, all *F*(2,106) >= 4.61, *p* <= .012, *f*_*p*_ >= 0.3, with the ERP responses being larger (i.e., more negative CNV and SPN, and more positive P3) for S than N (all *p*_*corr*_ <= .012, all *est* >= .61) and in SPN and P3 for S than M (all *p*_*corr*_ <= .04, all *est* >= .49). Additionally, in the reception, P3 was also statistically significantly larger for M than N (*p*_*corr*_ = .002, *est* = 0.59). Group was not a statistically significant predictor in these models (all *f*_*p*_ <= 0.18).

##### 3.1.1.3 Pupil sizes

Condition was a statistically significant predictor of pupil sizes in early anticipation, *F*(2,90) = 5.74, *p* = .004, *f*_*p*_ = 0.36, and in late anticipation, *F*(2,92) = 3.88, *p* = .024, *f*_*p*_ = 0.29 (in reception *f*_*p*_ = 0.19). In both anticipation phases pairwise post-hoc tests revealed that the main effect of condition was driven by larger dilations to S than N (all *p*_*corr*_ <= .035, *est* >= 0.04) and in early anticipation also to S than M (*p*_*corr*_ = .003, *est* = 0.05). Group did not significantly predict pupil size in any model (all *f*_*p*_ <= 0.1).

#### 3.1.2 Psychopathological approach (low AQ vs. ASC)

##### 3.1.2.1 Reaction times

The analysis of reaction times yielded a significant effect of condition, *F*(2,88) = 3.64, *p* = .03, *f*_*p*_ = 0.29, with faster responses in M than in S (*p*_*corr*_ = .037, *est* = 5.74). Group did not predict the responses, *f*_*p*_ = 0.04. For details, see sections 6.1.6. in the html file (and additionally 6.1.4.-6.1.5. for separate analyses of uncorrected reaction times and accuracy).

##### 3.1.2.2 ERPs

The models yielded a main effect of condition in early anticipation, *F*(2,104) = 5.25, *p* = .007, *f*_p_ = 0.32, with larger CNV to S (*p*_*corr*_ = .006, *est* = 0.45) and M (*p*_*corr*_ = .03, *est* = 0.36) in comparison to N, and in reception, *F*(2,104) = 20.79, *p* < .001, *f*_*p*_ = 0.63, with the largest P3 amplitudes to S, than to M, and smallest to N (all *p*_*corr*_ <= .004, all *est* >= 0.64). Condition did not predict the SPN amplitudes in late anticipation (*f*_*p*_ = 0.17). Group was a significant predictor only in early anticipation, *F*(1,52) = 4.83, *p* = .032, *f*_*p*_ = 0.31, where the CNV amplitudes were larger in the ASC than in the LAQ group (effect sizes for group in late anticipation and in reception were *f*_*p*_ <= 0.16).

##### 3.1.2.3 Pupil sizes

Condition predicted pupil sizes in early anticipation, *F*(2,84) = 5.57, *p* = .005, *f*_*p*_ = 0.36, and in late anticipation, *F*(2,82) = 5.63, *p* = .004, *f*_*p*_ = 0.37 (in reception *f*_*p*_ = 0.21). Pupil sizes were larger to S than N in both anticipation phases (all *p*_*corr*_ <= .033, *est* >= 0.04) and to S than M in early anticipation (*p*_*corr*_ <= .03, *est* >= 0.05). Group significantly predicted pupil sizes only in reception, *F*(1,42) = 7.27, *p* = .01, *f*_*p*_ =0.42, with larger constrictions in ASC than LAQ (in anticipation phases both *f*_*p*_ <= 0.17).

#### 3.1.3 Debriefing questions

We found no significant differences in self-reported general motivation during the experiment across autistic traits, *r*(79) = .05, *p* = .69, or between groups (both *t* <= 0.63, *p* >= .535). In contrast, the type of reward was reported to be more important for those with less autistic traits, *r*(79) = -.37, *p* < .001, and for LAQ than ASC (*t* = 3.5, *p* < .001; in LAQ vs. HAQ, *t* = -0.64).

Three participants reported that they never or almost never knew whether they were successful in the game directly after giving response, while the rest reported they knew sometimes (16), often (24), most of the time (33), or always (3). This did not differ significantly between the groups (both *p* >= .09).

Figure 3 displays average ratings of motivational values of the cues and of reward values of the feedback stimuli across groups. There was a statistically significant interaction of group and condition on subjective ratings of cues’ motivational values, *F*(4,152) = 2.77, *p* = .03. Although all groups showed descriptively higher ratings for the rewarded conditions (S and M) than N, post-hoc tests revealed that this was statistically significant in the LAQ and HAQ groups (all *est* >= 22.89, *p*_*corr*_ < .001) but not in the ASC group (all *est* >= 10.73). Moreover, LAQ on average rated S cues higher than ASC (*est* >= 23.96, *p*_*corr*_ = .015).

**Figure 3.**
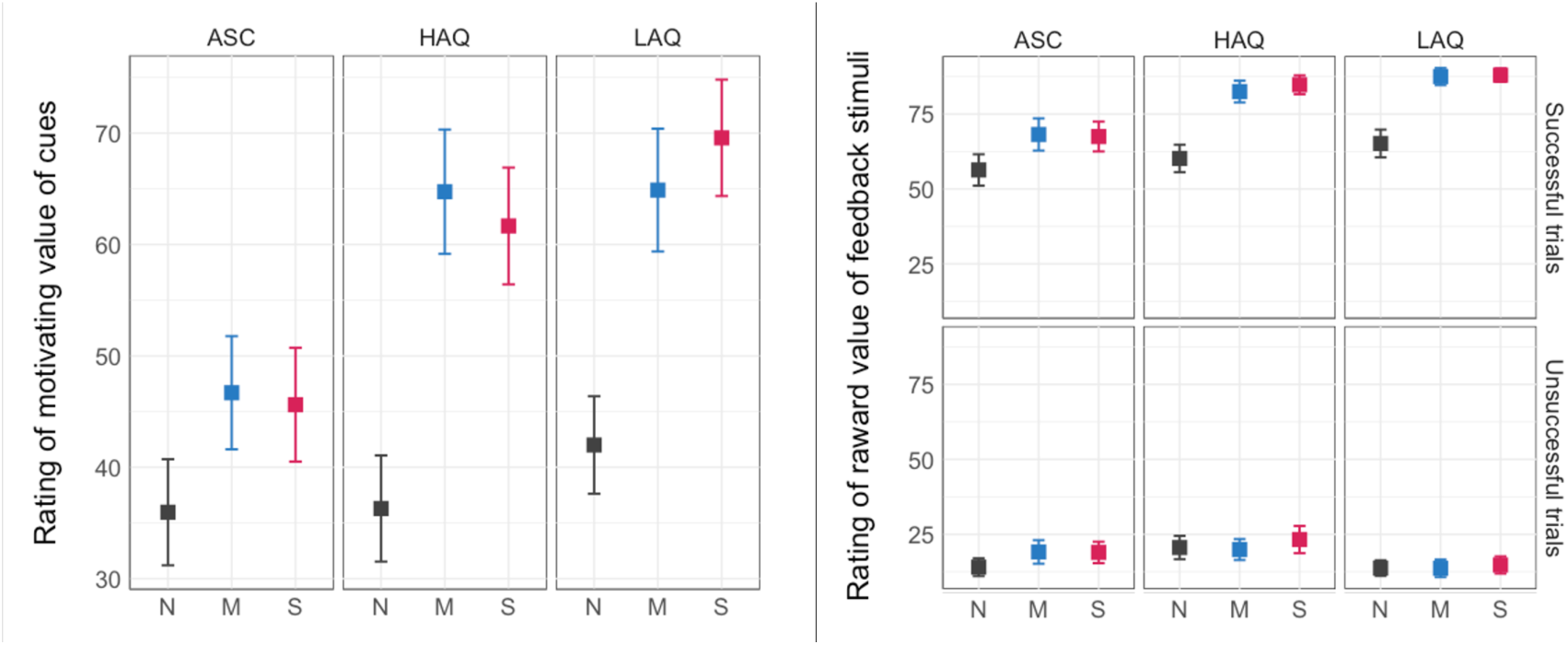
Average group ratings of (left) motivational value of the cues and (right) reward value of the feedback stimuli, separately for successful and unsuccessful outcomes.

For the analysis of subjective reward value of the feedback stimuli, we built a linear mixed model with group, condition, and outcome (successful or unsuccessful trial) as main predictors and with their interactions. This model yielded significant interactions of group and outcome, *F*(2,380) = 11.1, *p* < .001, and of condition and outcome, *F*(2,380) = 11.33, *p* < .001. Post-hoc tests revealed that the ratings of positive outcomes were higher in the non-autistic groups than in ASC (both *est* >= 11.81, *p*_*corr*_ <= .008) and, as expected, in all groups positive outcomes were rated higher than negative ones (all *est* >= 46.64, *p*_*corr*_ < .001). Feedback stimuli in all conditions were also rated higher in successful than unsuccessful trials (all *est* >= 44.39, *p*_*corr*_ < .001), and social and monetary rewards were rated higher than neutral positive outcomes (both *est* >= 18.89, *p*_*corr*_ < .001). For details and plots, see sections 4.6., 5.1.3., and 6.1.3. in the html file.

### 3.2 Secondary analyses

To summarise the highlights of the secondary analyses (see the supplementary material), we found that autistic traits were positively correlated with social anxiety traits and with stronger behavioural motivation to move away from unpleasant stimuli than to move towards desired outcomes (as assessed with the BIS/BAS). Further, the dimensional analyses (with AQ instead of group) paralleled the primary analyses: AQ did not interact with condition in either measure or phase, but higher AQ scores were linked to enhanced neuronal and pupillary responses respectively in early anticipation and in reception. Additional analyses revealed that the best fit for the relationship between AQ and all measures is linear, which suggests that autistic traits play a similar role in reward processing across individuals with and without autism. Finally, enhanced brain and pupillary responses were correlated across processing phases (early and late anticipation, reception), but not with each other.

## 4. Discussion

In this study, we investigated responsiveness to relevant social rewards, money, and neutral outcomes across autistic traits and in individuals with autism spectrum diagnosis, on multiple levels of processing. As hypothesised, using social stimuli relevant to the participants, we found that behavioural, neuronal, and autonomic responses of individuals with autism and higher levels of autistic traits were not differently influenced by the type of outcome (relevant social reward, money, neutral outcome) compared to the responses of subjects with low levels of autistic traits. However, individuals with autism, in contrast to those with low trait levels, showed enhanced reward responsiveness in early anticipation (larger CNV amplitude) and in reward reception (larger pupil constrictions). Both of these indexes have been previously linked to increased reward processing (Brunia et al., 2012; Cash-Padgett et al., 2018). These enhanced responses were also predicted by autistic traits across the whole sample in dimensional analyses. Finally, additional models revealed that the relationship between autistic traits and behavioural, neuronal, and autonomic responses to reward is likely linear.

In line with our hypotheses, we did not observe evidence for specific deficits in social reward processing in autism when using socially relevant stimuli. Models of all reward responses (neuronal, pupillary, and reaction times) yielded statistically insignificant interaction of group by condition, and using Bayes factors, we found strong evidence in favour of no interaction being the true effect in all the models. This stands at odds with the social motivation theory, which proposes that autism is characterised by diminished reward processing (i.e., hypo-responsiveness) specifically in the social domain (Dawson et al., 2002; Dawson, Webb, & McPartland, 2005; Schultz, 2005).

A potentially critical element in our design which could explain this is the relevance of the social rewards. While a common social stimulus in reward paradigms is a picture of a smiling, unknown person (e.g., Kohls et al., 2013; Scott-Van Zeeland et al., 2010; Stavropoulos & Carver, 2014b), in this study we used photographs of the main experimenter. The experimenter’s face became *familiar* to the participants during the study preparations, which they confirmed by recognising her in the photographs prior to the task. Importantly, the experimenter was also *socially relevant* in the context of the study, as she provided explanations and instructions, engaged in a semi-scripted, casual social exchange, and was present in the laboratory throughout the course of the study, also after the completion of the task by the participants. Therefore, while many previous studies used faces that were irrelevant in the study situation (even familiar faces, but absent or irrelevant in the context of the task; Neuhaus et al., 2015; Pankert et al., 2014; Stavropoulos & Carver, 2014a), we created a relevant social context. There is accumulating evidence suggesting that faces which are more familiar and relevant elicit higher activation in the brain reward structures (e.g., Acevedo et al., 2012; Bayer et al., 2021). Moreover, familiar, but not unfamiliar faces, have been reported to elicit neuronal and pupillary responses in ASC similar to those of comparison groups (Nuske et al., 2014; Pierce et al., 2004; Pierce & Redcay, 2008). Hence, we propose that the relevance of the social stimuli used in this study is an important qualitative factor which could have improved otherwise atypical responsiveness (as observed in other studies using unfamiliar faces, e.g., Kohls et al., 2011; Scott-Van Zeeland et al., 2010; Stavropoulos & Carver, 2014b) to social rewards in ASC and higher autistic traits.

Further, our data provide evidence that individuals with ASC, in comparison to those with low levels of autistic traits, show enhanced reward-related processing in the early anticipation and reception of rewards (indexed by increased amplitude of the CNV and larger pupil constrictions, respectively). While these results contradict accounts suggesting reduced responsiveness in ASC to social and non-social rewards (Bottini, 2018; Clements et al., 2018; Keifer et al., 2021; Kohls et al., 2012), they are not isolated in the literature. Autism has been repeatedly linked to enhanced neuronal activation in response to various rewards (Dichter et al., 2012; Groen et al., 2008; Pankert et al., 2014; van Dongen et al., 2015). Also, previous results from our group yielded a similar effect in early anticipation in subclinical levels of autistic traits (Matyjek et al., 2020).

This study investigated multiple levels of reward processing – neuronal, autonomic, and behavioural – with the aim to grasp a bigger picture of the process, which is necessary for an informed interpretation of predicted atypicalities in ASC. Across these levels, autism was linked to enhanced neuronal (early anticipation) and pupillary (reception) processing of rewards, but typical performance (reaction times) and decreased ratings of the motivational and rewarding values of the stimuli. One interpretation of these results is that individuals with ASC are more sensitive to rewards on the neuronal level (measured directly in the brain electrical activity and with pupil sizes as a proxy of the LC activity; Aston-Jones et al., 1999). The enhanced early processing in this group can be then reflecting the rapid formation of a representation of a reward and the initial anticipatory processes. However, this neuronal enhancement is weaker or absent in the later processing stages and is not translated to performance, which suggests that the motivational power of the incentive cues is not sufficient to modulate behaviour. Finally, increased autonomic measures in reward reception may indicate robust processing of the feedback (cf. with results from Baumeister et al., 2020, who observed hyperactivation of the ventral striatum during reward reception in over 200 ASC participants, although only at an uncorrected level). This, however, did not translate to a higher perceived rewarding value of the stimuli (as indicated by lower ratings of the positive feedback in ASC in our data).

Alternatively, these group differences may be an indicator of less efficient neuronal processing of rewards in ASC in the sense that larger activation is required to achieve similar performance. Importantly, the enhanced neuronal and autonomic processing was predicted by levels of autistic traits, which quantify manifestations of socio-communicative, attentional, and imaginal behaviours characteristic for ASC (Baron-Cohen et al., 2001). This suggests that the enhanced reward responsiveness on the neuronal and autonomic levels is linked to more pronounced autistic expressions on the behavioural level, which cuts through the borders of diagnostic groups. This speaks to the value of the dimensional analysis of autistic traits in addition to the coarse group differences based on the diagnostic cut-off. In this vein, by exploring autistic traits as continuously distributed in the population, we showed that reward processing atypicalities are likely linked to these traits in a linear manner: the higher the autistic traits, the larger the reward-related responses.

As expected, both brain and pupil responses were consistently larger to relevant social rewards compared to neutral outcomes and (less consistently) monetary rewards regardless of autistic traits and reward processing phases (early anticipation, late anticipation, reception). This corresponds to higher ratings of motivational and reward value of the rewards in comparison to the neutral outcomes. On the other hand, reaction times corrected for accuracy were faster in monetary trials than in social or neutral ones. Although this suggests higher motivational value of the monetary rewards on performance and of social rewards on psychophysiological measures, it has been reported before that reward magnitudes can predict subjective motivation and arousal, but not performance (Watanabe et al., 2019). Together, the larger responses in social and monetary than in neutral trials observed on multiple processing levels support that our paradigm was successful in capturing reward processing.

While more research is needed before firm conclusions can be drawn, our data suggest that the neuronal, autonomic, and behavioural indexes of reward processing reflect distinctive mechanisms and together offer a broader picture of this function. In line with this, ERPs and pupillary responses across conditions did not correlate with each other, but in each processing level (neuronal and autonomic), we observed consistent correlations between reward phases. The SPN was positively associated with the CNV (both negative ERPs) and negatively with the P3 values, which suggests that the larger the late anticipation, the larger both the early anticipation and the reception of rewards. The pupil sizes in the reception phase correlated negatively with the pupil sizes in early anticipation, which suggests that the larger the anticipation (indexed as increased dilations), the larger the reception (indexed as increased constrictions). These consistencies emphasise the additive explanatory values of ERPs and pupil sizes and emphasise the importance to investigate reward function on multiple levels.

The current study design, although based on a well-established paradigm (cued incentive delay task; Knutson et al., 2005) includes several aspects which allow us to disentangle potentially confounding factors in reward processing. Firstly, we used symbolic incentive cues which were not themselves rewarding (in contrast to showing a coin or a smiling face as a cue; Kohls et al., 2011). Thus, we ensured that the responses in early and late anticipation were indeed reflecting reward anticipation and not reception. Further, we included a non-rewarded condition (neutral), in which informative feedback was provided, but which did not offer any external rewards. Due to this, the observed enhanced responses to the social and non-social conditions in contrast to the neutral outcomes can be interpreted as reward processing on top of feedback processing. Finally, in all statistical models we controlled for social anxiety traits, as it is linked to atypical reward processing (Richey et al., 2014), and correlates with autistic traits (see point 1.1. in the supplementary material). This allowed us to interpret the obtained results as more autism-specific.

At the same time, several limitations in this study should be noted. First, we focused on adults but reward processing atypicalities linked to autism have been shown primarily in childhood (Kohls et al., 2011; Scott-Van Zeeland et al., 2010; Stavropoulos & Carver, 2014b) and are possibly dynamic throughout development (Keifer et al., 2021). In additional explorative analyses we observed that age was linked to diminished ERP responses in late anticipation and reception (see **Error! Reference source not found**. in the supplementary material). Similarly, exploratory models yielded that females exhibited increased pupil responses in late anticipation (dilations) than males (see **Error! Reference source not found**. in the supplementary material). However, it should be noted that groups in this study did not differ in age or gender distribution. Further, for the dimensional analyses we used the full AQ scores, but the social subscale of the AQ or the SRS would provide a more direct test of the social motivation theory’s predictions. This was not possible in our data, because many participants with ASC were not comfortable with the SRS, which needs to be filled out by a close person, and several provided a pre-existing full AQ score (from which we could not calculate the subscales’ scores). Finally, due to the need to maintain high experimental control over luminance and onset timing of the stimuli (for pupillometry and ERPs), we used static stimuli. Nevertheless, such stimuli are characterised by reduced ecological validity, especially in the social domain (Dziobek, 2012). Thus, future studies should attempt to replicate our results with dynamic stimuli.

To summarise, the present study provides evidence that autistic traits and autism are linked to atypical reward processing. However, in contrast to the social motivation theory, we observed enhanced neuronal and autonomic responses to both social and non-social rewards in autistic in contrast to non-autistic individuals with low levels of autistic traits. Importantly, we used social stimuli of relevance to the participants, which might have increased the reward value in the social condition and potentially mitigate otherwise atypical responsiveness to social rewards in autism and higher autistic traits. By investigating neuronal, autonomic, and behavioural responses, we provided a bigger picture of reward processing, which suggests a complex mechanism manifesting differently on each level. Autism in our data was linked to enhanced neuronal and autonomic processing, typical behavioural performance, and diminished self-rated responsiveness to rewards. We suggest that to understand reward responsiveness in autism, atypicalities found on the neuronal or autonomic levels must be interpreted in relation to the behavioural manifestations of social difficulties.

## Data Availability

Anonymised and aggregated data, analysis code in R, and an html file including all analyses steps and results can be found at https://osf.io/vse38/

## Acknowledgements

This work was supported by funding from the Berlin School of Mind and Brain, Humboldt-Universitaet zu Berlin and the German Academic Exchange Service (Deutsche Akademische Austauschdienst; DAAD) awarded to M.M. The authors are grateful to Sarah Margo Gawronska who helped with recruitment and data collection, and to the participants who volunteered to take part in the study.

## Supplementary material

**Table 1.** Means and standard deviations for ERP amplitudes, pupil responses, and reaction times in groups, conditions, and phases.

| Group | Condition | Early anticipation | Late anticipation | Reception |
| --- | --- | --- | --- | --- |
| ERPs [μV] | | | | |
|  |  | CNV | SPN | P3 |
| ASC | N | -0.32 (1.53) | -4.4 (3.42) | 2.98 (1.98) |
|  | M | -0.61 (1.77) | -4.34 (3.05) | 3.33 (1.89) |
|  | S | -0.49 (1.42) | -4.78 (3.1) | 4.23 (1.94) |
| HAQ | N | -0.21 (1.39) | -2.95 (3.34) | 1.73 (1.74) |
|  | M | -0.16 (1.71) | -2.97 (3.07) | 2.01 (1.83) |
|  | S | -0.67 (1.82) | -3.52 (3.47) | 2.81 (1.93) |
| LAQ | N | 0.33 (1.23) | -3.3 (2.81) | 2.23 (1.99) |
|  | M | -0.1 (1.17) | -3.53 (2.45) | 3.15 (2.16) |
|  | S | -0.41 (1.13) | -3.94 (2.71) | 3.82 (2.29) |
| Pupil sizes [au] |  |  |  |  |
| ASC | N | 0.11 (0.15) | 0.24 (0.26) | -0.49 (0.65) |
|  | M | 0.11 (0.14) | 0.23 (0.25) | -0.38 (0.55) |
|  | S | 0.17 (0.19) | 0.3 (0.25) | -0.4 (0.65) |
| HAQ | N | 0.05 (0.29) | 0.2 (0.23) | -0.19 (0.48) |
|  | M | 0.06 (0.26) | 0.22 (0.31) | -0.14 (0.5) |
|  | S | 0.07 (0.38) | 0.24 (0.36) | -0.15 (0.59) |
| LAQ | N | 0.08 (0.21) | 0.23 (0.29) | -0.1 (0.56) |
|  | M | 0.06 (0.17) | 0.28 (0.27) | -0.07 (0.6) |
|  | S | 0.1 (0.29) | 0.27 (0.25) | -0.17 (0.7) |
| Reaction times [ms] |  |  |  |  |
| ASC | N | 291.59 (49.65) |  |  |
|  | M | 292.79 (48.36) |  |  |
|  | S | 296.14 (44.97) |  |  |
| HAQ | N | 290.34 (50.79) |  |  |
|  | M | 270.16 (35.95) |  |  |
|  | S | 289.4 (62.21) |  |  |
| LAQ | N | 292.52 (41.73) |  |  |
|  | M | 283.48 (40.1) |  |  |
|  | S | 290.88 (39.83) |  |  |

## Exploratory analyses

All steps of the following analyses can be found in the html file available in the corresponding repository (https://osf.io/vse38/).

### 1.1 Questionnaires and brain-behaviour correlations

Correlations between the questionnaires, ERPs, and pupil responses were calculated using Pearson’s rank correlation coefficients.

#### 1.1.1 Questionnaires

We explored correlations between the AQ and other questionnaires and found statistically significant correlations for LSAS-SR, *r*(79) = .67, *p* < .001, BIS scale, *r*(79) = .45, *p* < .001, BAS drive scale, *r*(79) = -.23, *p* = .041, BAS fun seeking scale, *r*(79) = -.49, *p* < .001, and BAS reward responsiveness, *r*(79) = -.31, *p* = .006. For details and plots, see section 4.5. in the html file. Altogether, these results suggest that higher autistic traits are related to decreased social ability, increased social anxiety, higher sensitivity of the inhibition system, and reduced activation of the approach system. Thus, these data support that individuals with high autistic traits have stronger behavioural motivation to move away from unpleasant stimuli than to move towards desired outcomes.

#### 1.1.2 Correlations of the brain and pupil responses

Across all successful trials, the mean SPN correlated positively with the mean CNV, *r*(79)= .27, *p* = .018, and negatively with the P3, *r*(79) = -.58, *p* < .001. This suggests that larger brain responses in late anticipation are linked to also larger responses in early anticipation and in reception.

The mean pupil sizes were negatively correlated in reception and in early anticipation, *r*(66) = -.54, *p* < .001. This suggests that larger pupil responses in anticipation (i.e., more dilation) are related to larger pupil responses in reception (i.e., more constriction).

We found no significant correlations between ERPs and pupil sizes. Figure 1 shows correlogram of brain and pupillary responses. For details, see section 7.4. in the html file.

**Figure 1.**
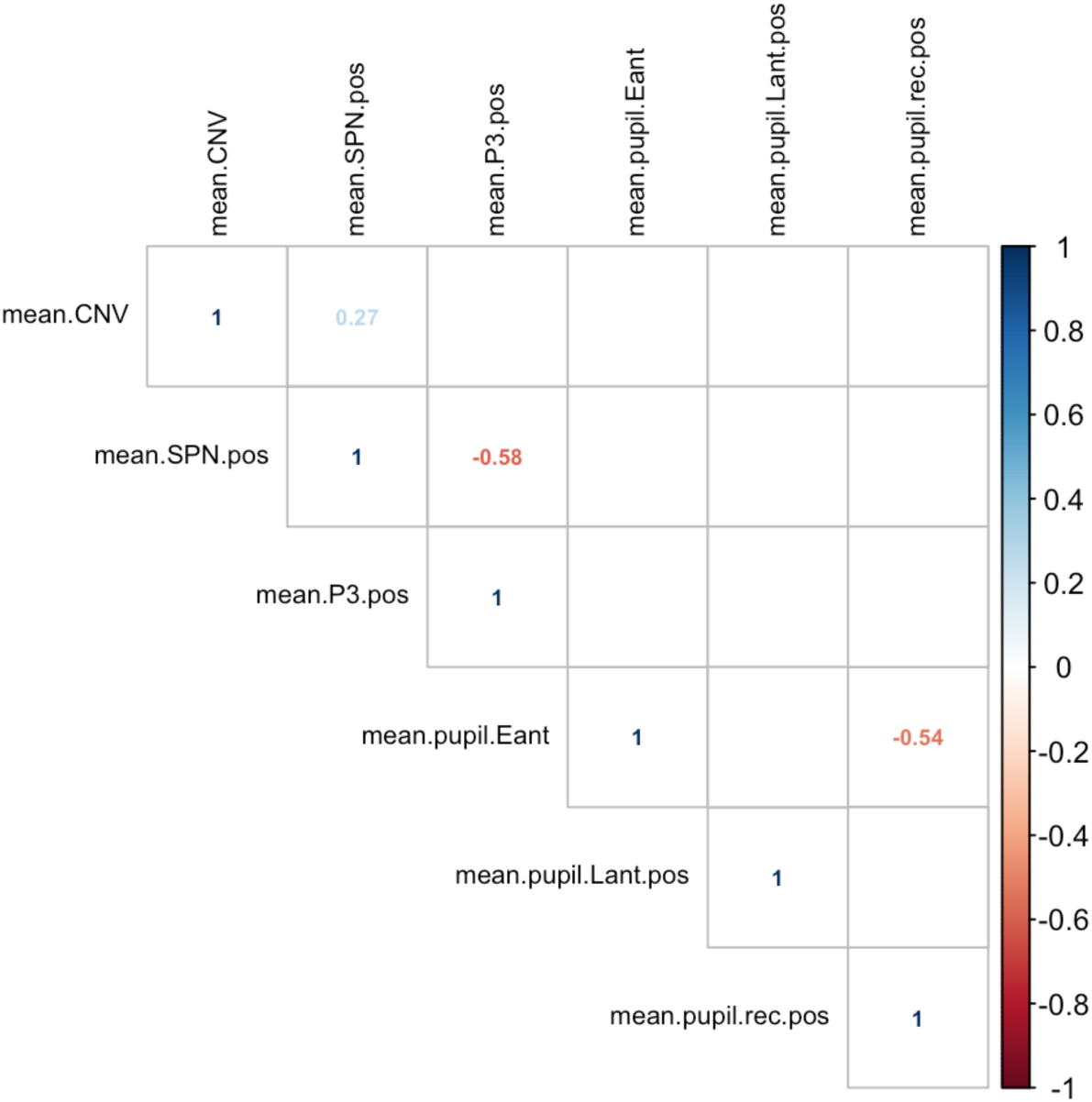
Correlogram: ERPs and pupil sizes in successful trials across conditions. Coefficients are displayed only for statistically significant correlations.

#### 1.1.3 Correlations of brain, pupil, and self-reported data

The mean anticipatory brain responses (but not the P3) across conditions were found to correlate with the debriefing questions and questionnaires: Higher self-reported general motivation in the experiment was linked to larger CNV amplitudes, *r*(79) = -0.26, *p* = .019, higher BIS scores were linked to larger SPN amplitudes (in successful and unsuccessful trials, both *r*(79) >= -0.37, *p* <= .005), and higher BAS fun seeking scores in unsuccessful trials were linked to smaller SPN amplitudes, *r*(79) = 0.27, *p* = .017.

The mean pupil size across conditions correlated with self-reported importance of condition, so that the more important the condition, the weaker the pupil response in early anticipation and in reception of unsuccessful feedback (smaller pupil size in early anticipation, i.e., weaker dilations, and larger pupil size in reception, i.e., weaker constrictions), respectively *r*(66) = -0.25, *p* = .04 and *r*(66) = 0.34, *p* = .005. For details, see section 7.3. in the html file.

### 1.2 Effects of group, condition, and outcome (successful and unsuccessful trials) on ERP and pupillary responses in the reception phase

To explore differences between reception of successful and unsuccessful outcomes on the neuronal and pupillary responses, we built models including group, condition, outcome (successful and unsuccessful), and their interactions. The predicted P3 and pupillary responses in successful and unsuccessful trials across all groups are shown in Figure 2 and Figure 3. Full analyses can be found in section 7.2. of the html file.

**Figure 2.**
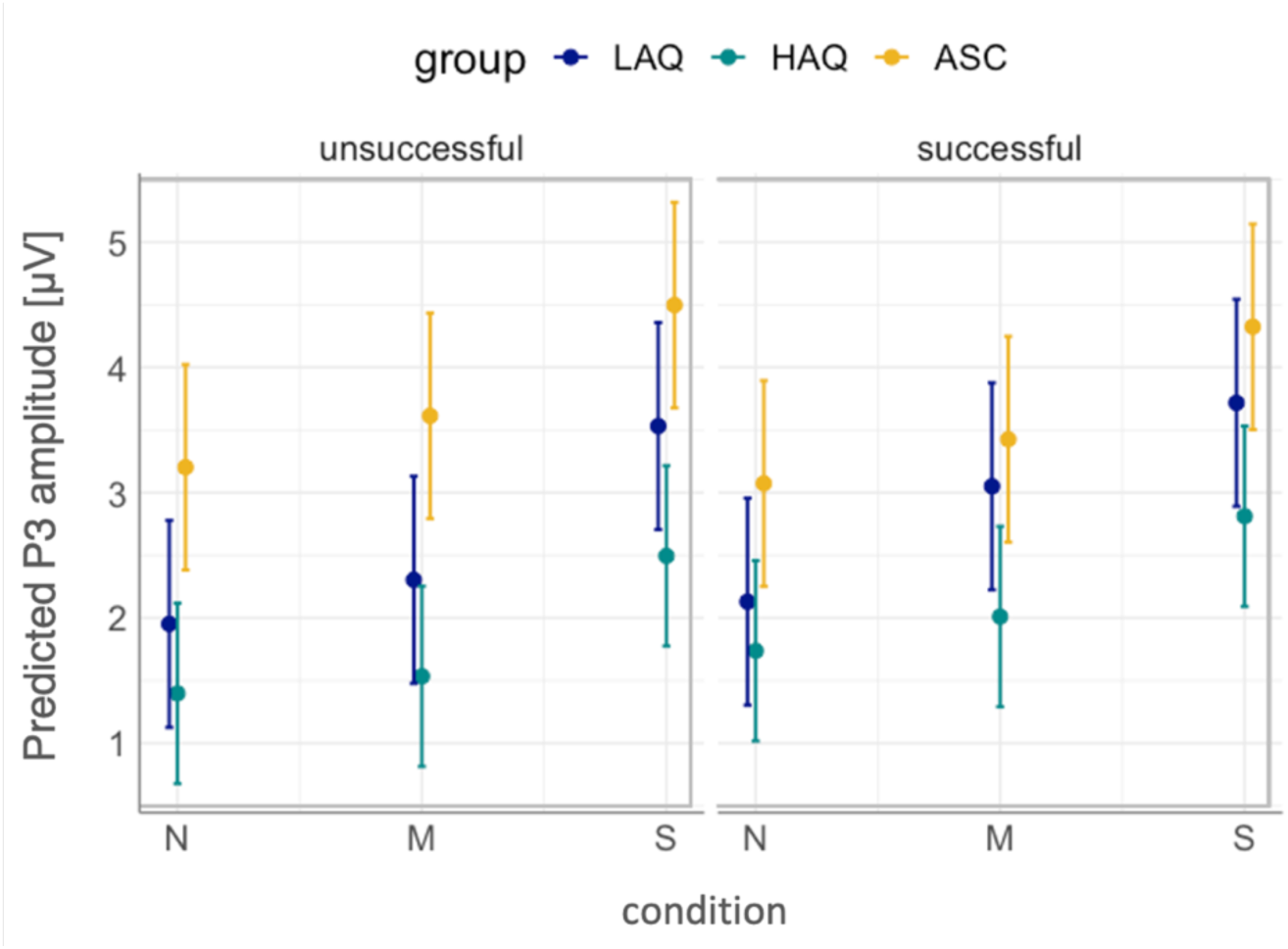
Average P3 responses in successful and unsuccessful trials across groups.

**Figure 3.**
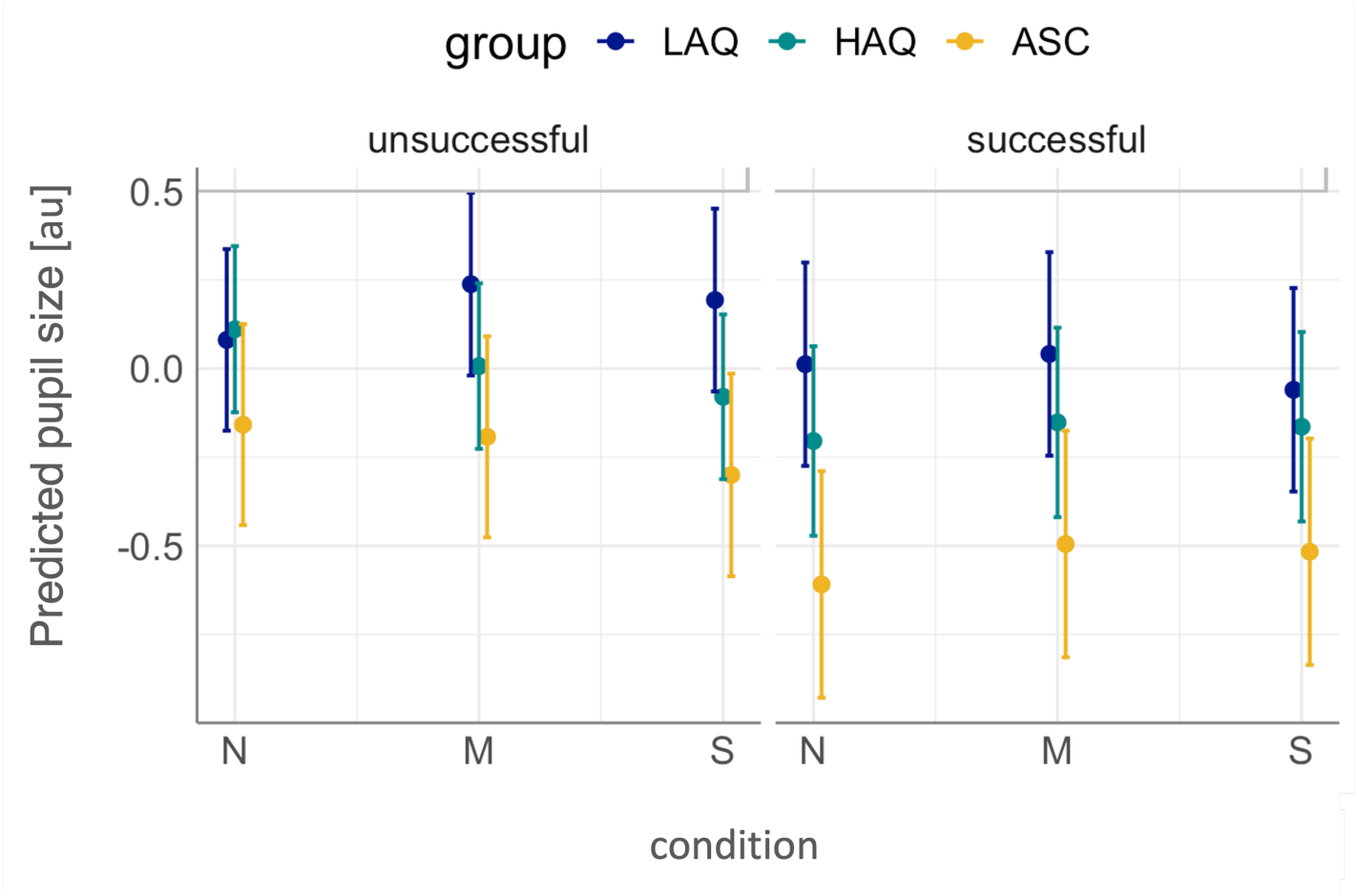
Average pupil sizes in successful and unsuccessful trials across groups

For neuronal responses, we observed an interaction of group and outcome (*F*(2,395) = 3.53, *p* = .03, *f*_*p*_ = 0.13). This was driven by larger P3 amplitudes in ASC as compared to HAQ, for both successful and unsuccessful trials (both *p*_*corr*_ <= .034). The pupillary data revealed significantly larger pupil constrictions to outcomes in successful than in unsuccessful trials, *F*(1,513) = 31.24, *p*_*corr*_ < .001, *f*_*p*_ = 0.25 (no other contrasts were significant).

### 1.3 Dimensional analyses – AQ as a predictor of reaction times, ERPs, and pupillary responses

In addition to the pre-registered analyses, we explored the effects of autistic traits across the sample on reward responses (a dimension analysis). Because in the spectrum view of autism the distribution of autistic traits in the general population is continuous, we conducted exploratory analyses in which the main predictor of reward responsiveness is continuous AQ instead of group. Finally, it is conceptually interesting to consider whether the potential atypicalities in reward processing would increase linearly with higher autistic traits in the population, or whether this increase would become steeper with particularly high trait levels (for example, around the approximate cut-off of autism diagnosis). To investigate this, we also built exploratory generalised additive mixed models (GAMMs) with package mgcv ver. 1.8-31 (Wood, 2011) in the following form:

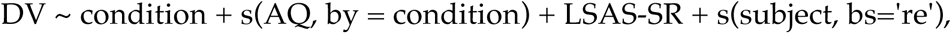

where DV is the dependent variable, s(AQ, by = condition) is a smooth term for AQ fitted separately for each condition, and s(subject, bs=‘re’) is the random smooth for subjects. AQ and LSAS-SR were centred before they entered the statistical models. Here, we report only the main effects of those additional models and the complete analyses can be found in the analysis code in the referred repository. Figure 4 shows predicted neuronal and pupillary responses across levels of autistic traits in all phases. All analysis steps are shown in section 7.1. of the html file.

**Figure 4.**
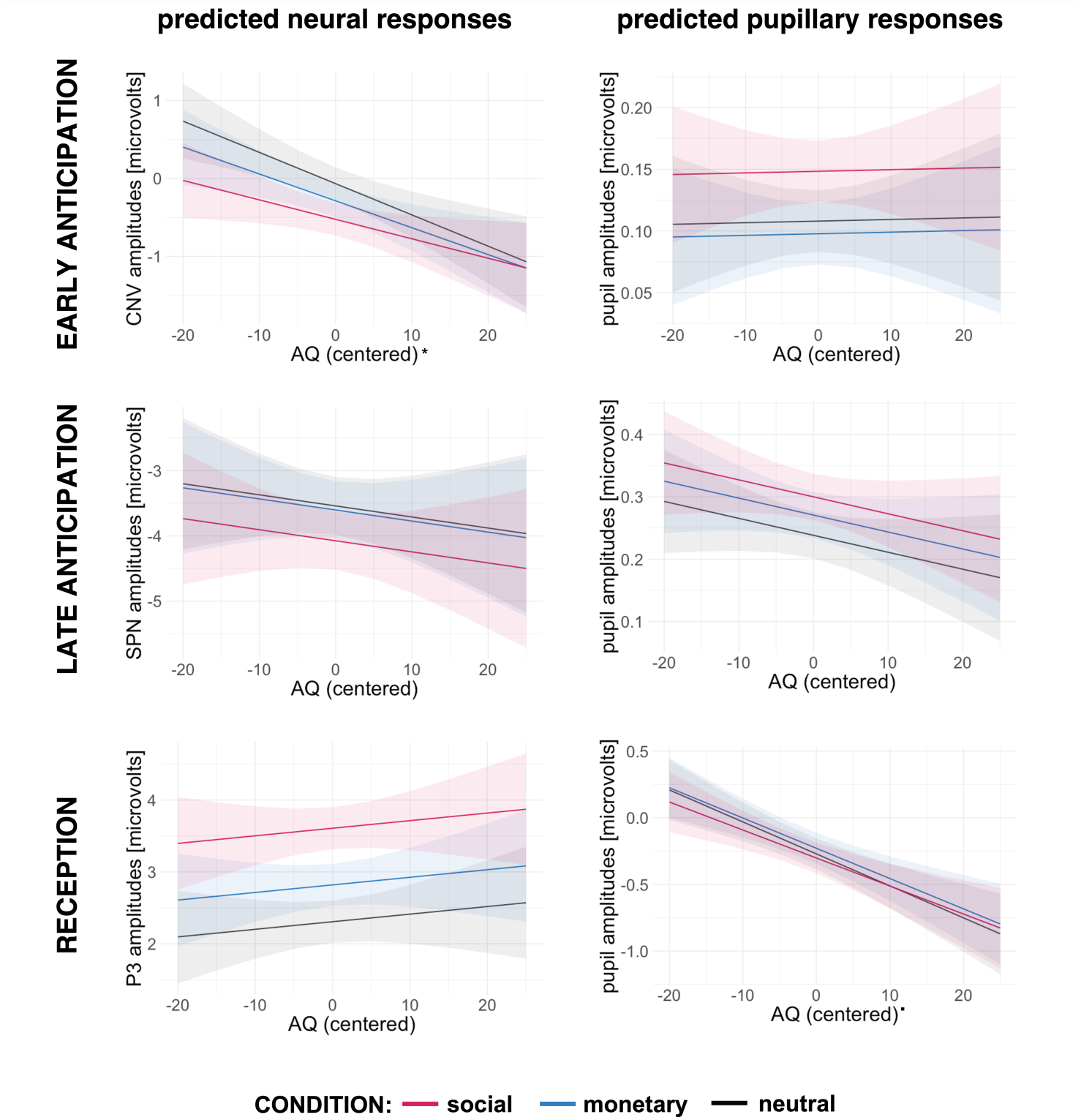
Predicted values of neuronal and pupillary responses to social, monetary, and neutral outcomes in successful trials across autistic traits (AQ) in all participants. Shadowed areas represent 95% confidence intervals. Statistically significant effects of AQ were marked with * for p < .05

#### 1.3.1 ERPs

Condition was a significant predictor of the brain responses in all linear models with continuous AQ (instead of group): early anticipation, *F*(2,158) = 7.76, *p* = .001, *f*_*p*_ = 0.31, late anticipation, *F*(2,156) = 3.57, *p* = .031, *f*_*p*_ = 0.21, and reception, *F*(2,158) = 4.32, *p* = .015, *f*_*p*_ = 0.23. In all models, responses to S were larger than to N (all *p*_*corr*_ <= .031, *est* >= 0.46). Additionally, in late anticipation and reception, S elicited larger ERP amplitudes than M (both *p*_*corr*_ <= .035, *est* >= 0.47), and in reception M was linked to larger P3 than N (*p*_*corr*_ =.002, *est* = 0.51). The AQ score significantly predicted the brain responses only in the early anticipation, *F*(1,79) = 4.28, *p* = .042, *f*_*p*_ = 0.23 (in late anticipation and reception *f*_*p*_ = 0.05), with higher AQ scores linked to larger (more negative) CNV response.

GAMMs with continuous AQ score yielded similar pattern of effects: condition (entered as a parametrical term) significantly predicted ERP amplitudes in all models (all *F* >= 4.26, *p* <= .012) with larger responses to S than N. In the early anticipation model (and not late anticipation and reception), the AQ smooth was significant (*F* = 4.12, *p* = .044). Importantly, in all models AQ was fitted with effective degrees of freedom (edfs) of 1 (in reception edf = 1.2), which suggests that the best approximation of the relationship between autistic traits and reward-related brain responses is linear.

#### 1.3.2 Pupil sizes

In both anticipation phases, the pupil sizes were predicted significantly by condition: early anticipation, *F*(2,128) = 9.66, *p* < .001, *f*_*p*_ *>*= 0.39, and late anticipation, *F*(2,128) = 5.66, *p* = .004, *f*_*p*_ = 0.29. Responses were larger in S than in N (both *p*_*corr*_ <= .018, *est* >= 0.04) and in early anticipation also in S than M (*p*_*corr*_ < .001, *est* >= 0.05). AQ approached significance in reception, *F*(1,42) = 3.33, *p* = .075, *f*_*p*_ = 0.33 (in anticipation phases both *f*_*p*_ <= 0.29).

GLMMs yielded a main effect of condition in both anticipatory phases (both *F* >= 5.41, *p* <= .005) with larger responses (more dilation) to S than N. In all phases, the models fit better without separate AQ smooths for conditions and with edf = 1. AQ was statistically significant only in reception with higher AQ scores linked to smaller pupil sizes (*F* = 6.44, *p* = .012).

#### 1.3.3 Reaction times corrected for accuracy

Condition was a significant predictor of the LISAS scores, *F*(2,136) = 8.6, *p* < .001, *f*_*p*_ = 0.36, with faster responses to M than to S and N (both *p*_*corr*_ <= .001, *est* >= 6.31). The AQ did not significantly predict LISAS (*f*_*p*_ = 0.04). Figure 5 shows predicted LISAS scores across AQ.

**Figure 5.**
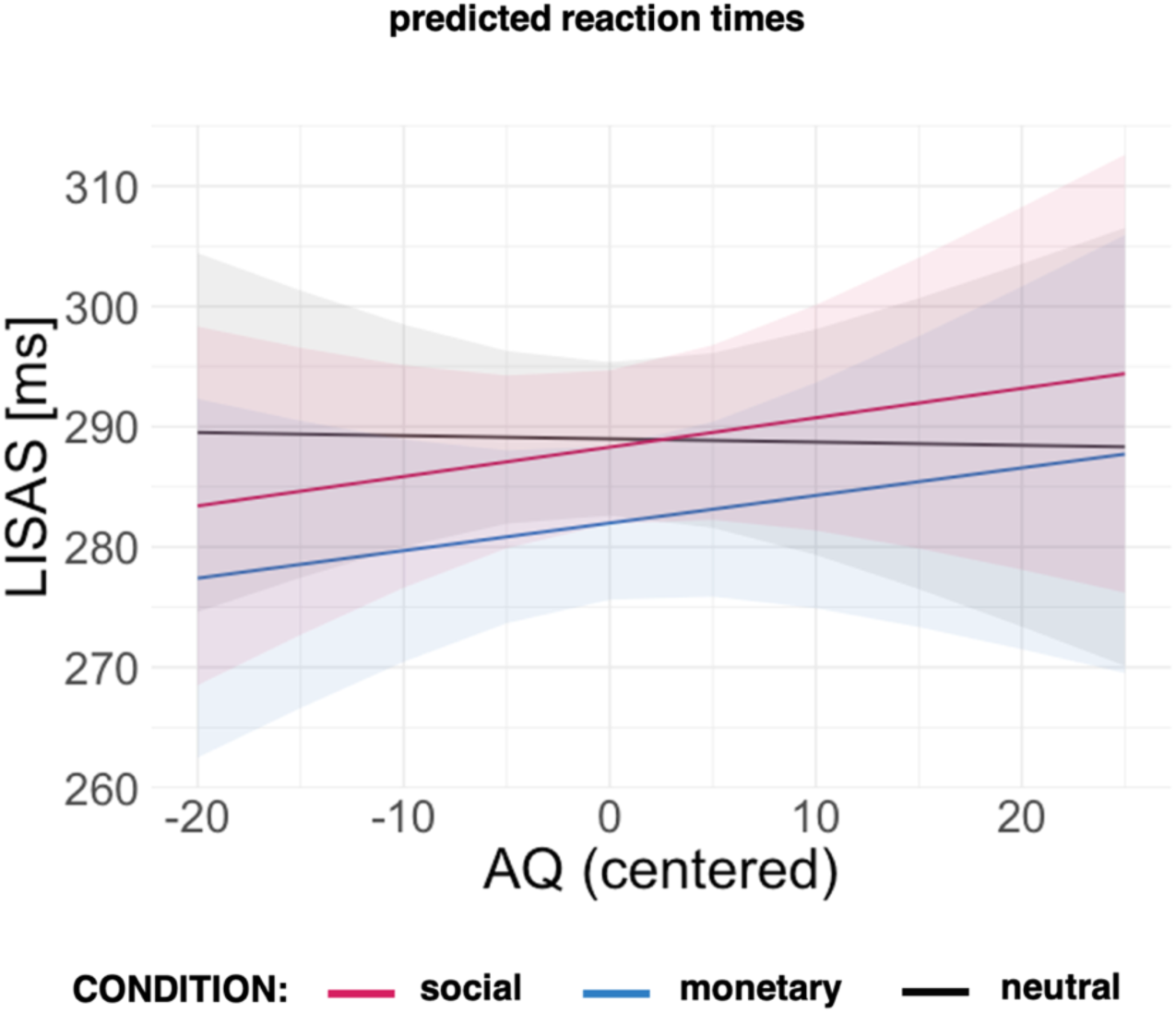
Predicted reaction times corrected for accuracy in social, monetary, and neutral conditions in successful trials across autistic traits (AQ) in all participants. Shadowed areas represent 95% confidence intervals.

The GAMM model showed a slightly better fit for a separate smooth for AQ in each condition, with edf = 1 for N and M and 1.3 for S. However, none of the smooths were significant. Condition as a parametric term significantly predicted LISAS (*F* = 8.62, *p* < .001), with the fastest responses in M.

### 1.4 Age and gender effects in ERP and pupillary models

For all primary models, we additionally explored the effects of age and gender. New models were in the form:

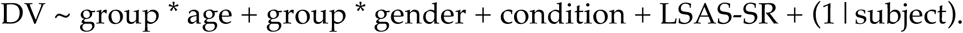

**Figure 6.**
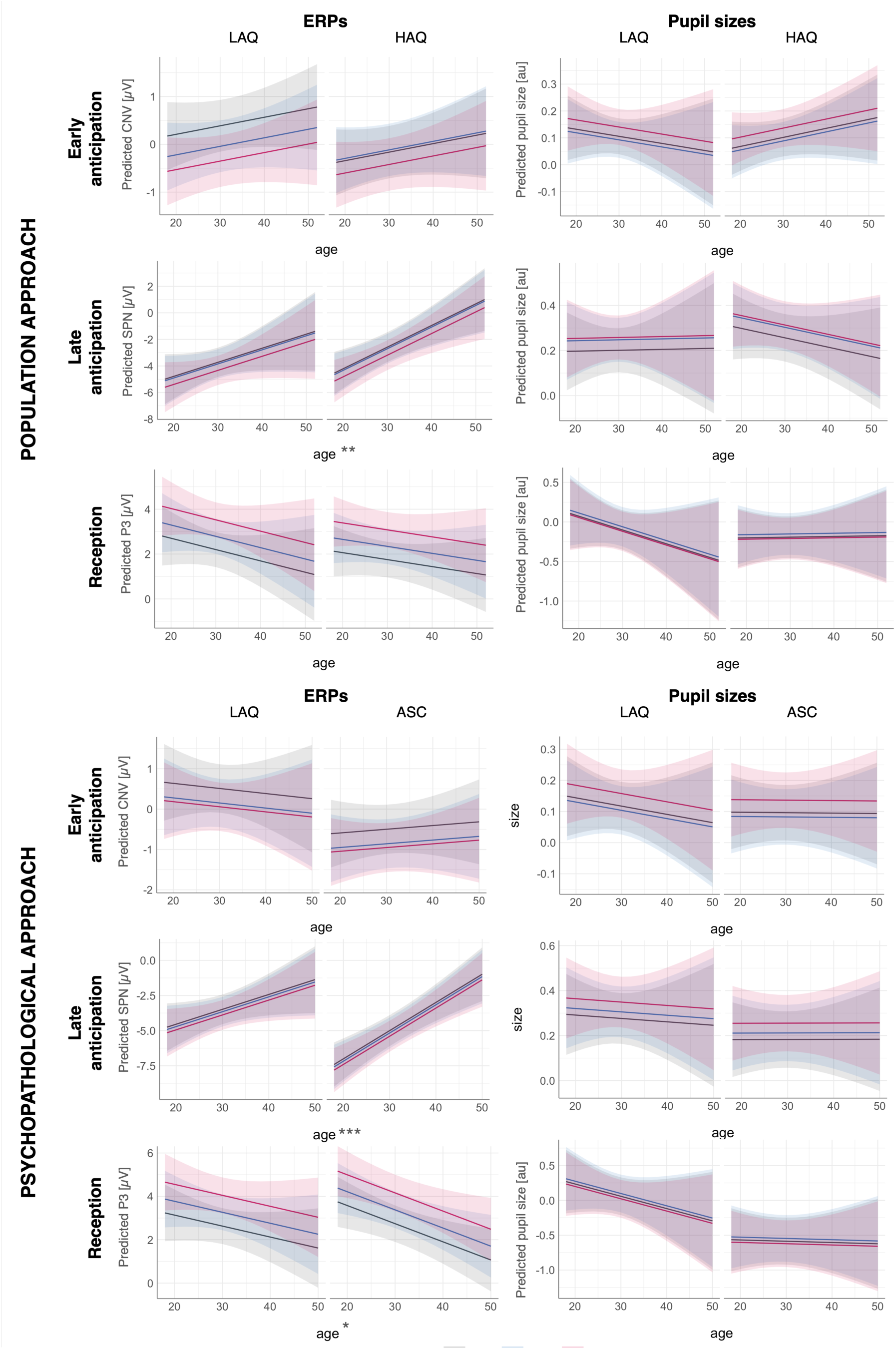
Effects of age on ERP and pupillary responses across groups (LAQ, HAQ, ASC), conditions (N = neutral, M = monetary, S = social), and phases (early/late anticipation, reception). Statistically significant predictors are marked with * for p<.05, ** for p<.01, and *** for p<.001. Details and full analyses can be found in the html file in sections 5.2-5.3 and 6.2-6.3.

**Figure 7.**
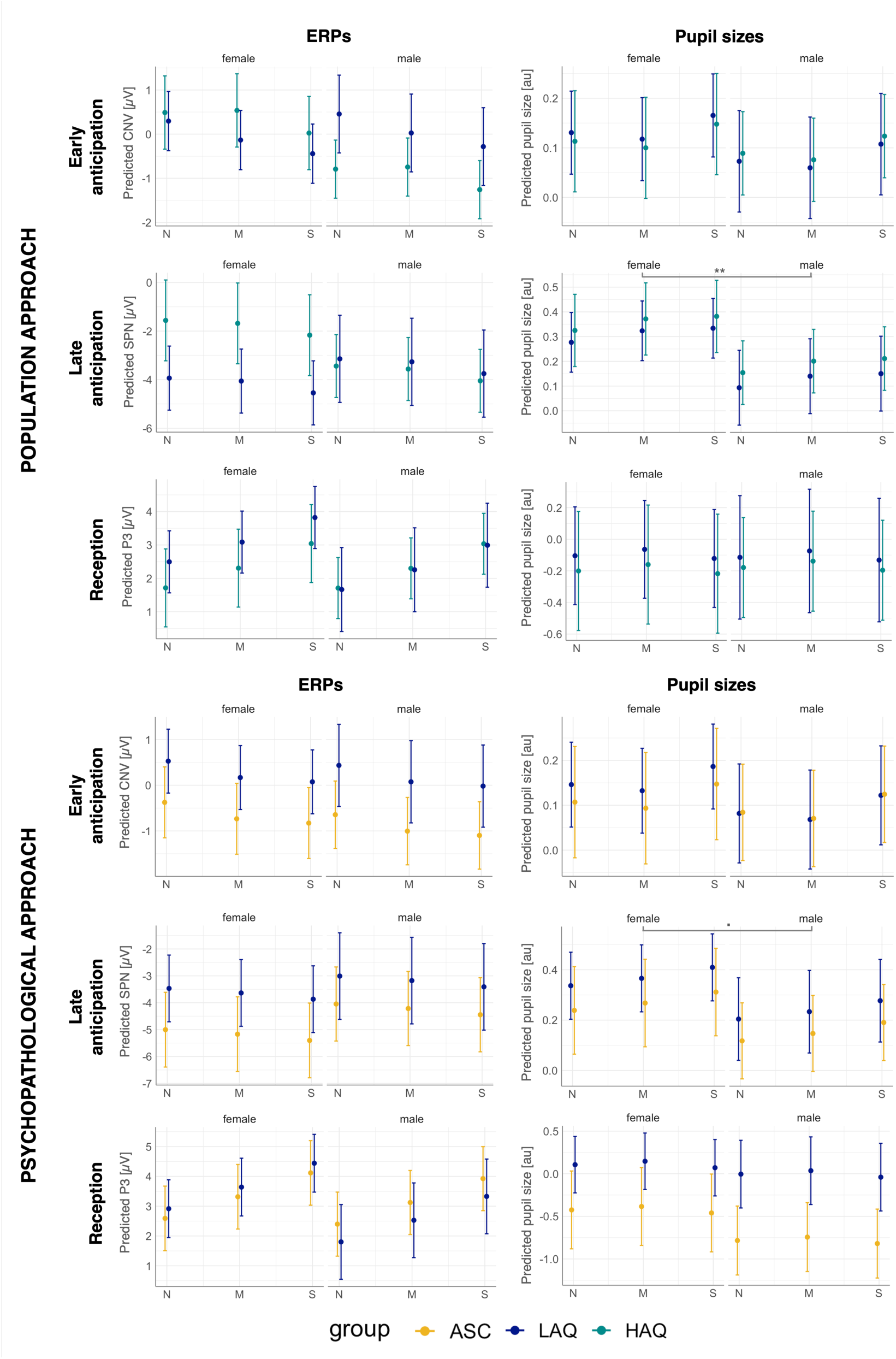
Effects of gender on ERP and pupillary responses across groups (LAQ, HAQ, ASC), conditions (N = neutral, M = monetary, S = social), and phases (early/late anticipation, reception). Statistically significant predictors are marked with * for p<.05, ** for p<.01, and *** for p<.001. Details and full analyses can be found in the html file in sections 5.2-5.3 and 6.2-6.3.

